# Non-Medical COVID-19 Impacts and Hearing Status: A Global Study of Differential Health Impact Among Deaf, Hard of Hearing, and Hearing Populations

**DOI:** 10.64898/2026.06.09.26355192

**Authors:** Shazia Siddiqi, Madeline Murray, Wyatte C. Hall, Michelle Koplitz, Timothy De Ver Dye

## Abstract

**Background:** Deaf and hard-of-hearing (HoH) experienced complex challenges during the COVID-19 pandemic, including obscured visual communication from mask mandates, inaccessible public health messaging, and inadequate interpreter availability. We examined whether hearing status predicted non-medical COVID-19 impact on a global level.

**Methods:** We conducted a nested cross-sectional analysis within a global study collecting data across two waves (April–May 2020 and July–August 2022) from 184 countries. Participants (N=7,998) were categorized as Deaf (n=304), Hard of Hearing (HoH; n=951), or Hearing (n=6,743). The primary outcome was a composite COVID-related non-medical Personal Impact T-Score derived from 14 items across employment, resource access, and healthcare domains. Multinomial logistic regression models progressively adjusted for demographic, structural, and psychosocial variables.

**Results:** Deaf participants reported substantially higher rates of pandemic-related job loss (28.9% vs. 9.6% hearing), healthcare cancellations (39.9% vs. 24.6%), and inability to obtain basic supplies. Over half (55.9%) of Deaf participants scored above the median composite impact index, compared to 39.2% of hearing participants. In the fully adjusted model, Deaf status remained an independent predictor of high non-medical impact (aOR=1.6, 95% CI: 1.1–2.4). HoH status showed no statistically significant difference from hearing participants in any model.

**Conclusions:** People identifying as Deaf experienced significant disparities during COVID-19 when compared with HoH or hearing people, driven by language access barriers and institutional exclusion rather than hearing loss per se. These experiences underscore the importance for systemic interventions centering on accessible communication, Deaf-centered needs, and reducing audism in Deaf-hearing interaction.

**Strengths and limitations of this study:**

- Data were collected across two pandemic waves (April–May 2020 and July–August 2022) from 184 countries, allowing us to analyze hearing status on a global level, beyond any single national context or health system.
- The survey was deployed in multiple spoken languages and in American Sign Language (video) alongside written English, and the investigator team included Deaf researchers, supporting direct and accessible participation by Deaf communities rather than reliance on hearing-mediated proxies.
- The composite non-medical impact score was constructed using an established psychometric method with a defined item-retention criterion, and multinomial regression models were progressively adjusted for demographic, structural, and psychosocial variables.
- Participants were recruited through non-probability convenience sampling via social media and Amazon Mechanical Turk, so the sample is not population-representative and is subject to self-selection and coverage bias.
- Hearing status was self-reported and several distinct self-identified categories were consolidated into three groups (Deaf, Hard of Hearing, Hearing), which could mask heterogeneity within these identities.

## Introduction

The COVID-19 pandemic disproportionately affected marginalized and vulnerable populations worldwide, exacerbating known and revealing other systemic inequities that expanded beyond medical outcomes. While the clinical implications of the SARS-CoV-2 infection have been well-documented, the non-medical consequences of the pandemic have not been sufficiently addressed, especially in people with disabilities. Among these populations, deaf and hard-of-hearing (DHH) individuals faced socioeconomic, educational, and healthcare challenges in particular during the pandemic.

The public health response created additional major barriers for the DHH population. For instance, face mask mandates, essential for infection control, obscured visual facial cues critical for lipreading and sign language comprehension, causing communication difficulties, social isolation, and reduced well-being.^1,2^ DHH individuals also encountered systemic difficulty obtaining timely, accurate, and trustworthy health information in accessible formats, including their native sign languages. Indeed, DHH adults in the U.S. reported knowledge of COVID-19 symptoms comparable to that of hearing peers, but were also 4.7 times more likely to report difficulty accessing COVID-19 information and heightened mistrust in public health information when it was not available in American Sign Language.^3^ DHH individuals with limited health literacy were at higher risk of information marginalization during the pandemic.^4^ This information marginalization widened health literacy gaps and forced many to rely on less credible sources, such as social media, heightening their vulnerability.^5^ Similar inequities were documented internationally among older DHH individuals in China who received suboptimal medical access during emergencies,^6^ and within Deaf communities in Italy, where supportive social and cultural networks were needed to foster health literacy and advocacy in the face of systemic barriers.^7^

These communication barriers have cumulative effects on the DHH population’s daily lives. In the U.S., studies found that DHH adults reported alarming rates of food insecurity (42%) and isolation-related distress (54%) during the early restrictions of the pandemic.^8^ In Australia, parents of DHH children documented significant declines in socio-emotional well-being and challenges managing distance learning demands during the first year of the pandemic.^9^ These educational barriers further compound longstanding communication access inequities that predispose DHH populations to health disparities. The expansion of telehealth, which reshaped the healthcare delivery landscape throughout the pandemic, did not meet the accessibility needs of DHH individuals due to limited interpreter availability, providers’ limited awareness of telehealth systems, and inadequate cultural competence among providers.^10,11^ Even COVID-19 vaccination rates were lower among adults with hearing loss than among their non-disabled peers.^12^

Globally, the pandemic negatively affected health-related quality of life (HRQOL), with factors such as age, sex, education level, health status, and economic security influencing individuals’ HRQOL.^13^ As a result, those who faced systematic barriers due to these aspects were at higher risk of a decline in their quality of life. For DHH individuals who experienced new challenges due to pandemic constraints (e.g., masking, telehealth, and access to interpreters), these challenges exacerbated inequalities, leaving them more vulnerable to negative impacts on their overall quality of life.

The medical impact of COVID-19 on individuals and societies is substantial and clear.^14^ The differential cumulative social and lifestyle impacts of COVID-19, while creating morbidity and lasting consequences for individuals, their families, and communities, are less clear. As such, this study aimed to examine how hearing status, specifically that of those who are deaf and hard-of-hearing, related to non-medical COVID-19 impacts at a global level.

## Methods

### Study Design

We conducted a global nested analytical cross-sectional study assessing health status, psychosocial dynamics, and COVID-19 lived experiences, collected in two waves: during early COVID (April-May 2020), and later in the pandemic (July-August 2022)). Our focus in this nested analysis within the larger global study was to evaluate whether cumulative non-medical COVID-19 experiences differed among (1) Deaf, (2) Hard of Hearing (HoH), and (3) hearing people after controlling for mediating or confounding variables. This analysis would help us better understand not only hearing-related disparities but also evaluate the contribution of hearing status to lived experience during the pandemic.

The larger global study was implemented using the Critical Medical Ecological theoretical model as a guide, incorporating biological, social, health care, environment data, and power dynamics across the domains of individuals, households, and communities in six world regions (Africa, Asia, Europe, Latin America and the Caribbean, Northern America, and Oceania).^15^

We present the results in accordance with the STROBE checklist for cross-sectional studies.^16^

### Setting

We aimed for a global study that would help us assess variation in non-medical COVID-19 impact around the world with the understanding that the pandemic moved differently from country to country and – as a powerful infectious virus – interacted with ecosystems, cultures, and health systems that could impact the disease trajectories of communities. To achieve maximum reach, we deployed the survey through the REDCap survey platform in Spanish, English, French, and Italian during Wave 1, and in those languages, plus Mandarin Chinese, Arabic, and Hindi, during Wave 2. We also deployed the survey in American Sign Language (video) and in written English during the recruitment of Deaf communities. REDCap as a tool enabled the implementation of our survey in all these languages and modalities.

We recruited participants for this study from two main sources: social media during all waves of the survey (Facebook, Instagram, and the Facebook Audience Network) and (during Wave 1 only) through Amazon’s on-demand, scalable “Mechanical Turk” (mTURK) workforce. We used boost functions to distribute the survey throughout the world where the targeted languages were spoken to ensure the widest geographic coverage possible. Potential participants clicking on the invitation were routed to a REDCap (v. 9.9.2, Vanderbilt University) installation where they were presented with further details and an IRB-approved Information Sheet in one of the languages of the study. Participants were required to confirm that they were 18 or older and to enter their country of residence before proceeding with the rest of the online survey. Personal identifying details were not collected in the survey. Additional details about participants from the parent study are available elsewhere.^17^

### Variables

*Hearing status*.

Given that the investigator team included Deaf researchers with expertise in Deaf health research, we deliberately included self-identified hearing status as a variable in all survey versions. Self-identification as d/Deaf is complex, encompassing both physical and cultural aspects of the d/Deaf experience. In Wave 1, we first presented participants with an explanation of this variable, in each language:

*For the purposes of this study, please use the following definitions*.
**Hearing/not deaf**: Person with no hearing loss;
**Hard of hearing**: Person with some hearing loss;
**deaf**: Person that has hearing loss;
**Deaf**: Person that has hearing loss and identifies with Deaf culture;
**DeafBlind**: Person with a combination of hearing loss and limited-to-no vision.

We then asked participants: *Which of the following categories do you identify as?* And presented participants with the five options as described above.

In Wave 2, we added two additional categories: “Unilaterally deaf” and “having a hearing loss” based on feedback from participants and Deaf investigators.

Deaf people often self-identify as “Deaf” (with a capital D), which insinuates they consider themselves part of Deaf culture (typically including use of a signed language and identifying with elements of sociocultural Deaf norms).^18^ That said, deaf people who do not identify particularly with deaf culture, or who prefer not to self-identify as capital “D” Deaf, were able to select “deaf” with a lowercase “d.” Including DeafBlind, unilaterally deaf, and “having a hearing loss” helps participants self-identify with the group that best describes their affinity. For our study, we consolidated “Deaf,” “deaf,” and “DeafBlind” into one category (“Deaf”), “hard of hearing,” “unilaterally deaf,” or “having a hearing loss” as “Hard of Hearing,” and “Hearing/not deaf” as “Hearing.” We acknowledge these reductions may oversimplify or mask true heterogeneity in these identities.

#### Outcome: COVID-related non-medical Personal Impact Score and Subscores

We were guided by Streiner et al.’s method^19^ for constructing the main outcome of this analysis: the COVID-related non-medical Personal Impact Score. Items assessed for inclusion were binary (yes/no) responses to the following question (items marked with (*) were added by the research team; all other items originated with the Kaiser Family Foundation, KFF Coronavirus Poll March 2020): “Please tell us if you have taken any of the following actions because of the recent coronavirus outbreak” (Decided not to travel or changed travel plans; Bought or worn a protective mask; Stocked up on items such as food and household supplies; Postponed or canceled health care visits;* Got extra refills on prescription medication;* Stayed home instead of going to work, school, or other regular activities; Postponed or canceled a medical procedure or surgery;* Canceled plans to attend large gatherings such as concerts or sporting events; Quit my job*) in addition to yes/no responses to “Have you experienced any of the following because of coronavirus?” (Lost income from a job or business; Been unable to get groceries; Been unable to get cleaning supplies or hand sanitizer; Been unable to get prescription medication; Have you or a family member been harassed, bullied, or hurt because of coronavirus*). Items achieving 0.2 or higher corrected item-total correlation were retained in the Impact Score (all 14 items met this criterion), and the overall standardized Cronbach’s alpha for the resulting score was 0.65. To adequately account for item completion, the final Impact Score constructed for analysis averaged individual participant responses (number of “yes” responses to items answered divided by the total number of items answered) for respondents who answered at least half (7) of the individual impact items. The raw mean was converted to a z-score, then transformed to a standardized T-score with a mean of 50 and a standard deviation of 10. The resulting score was the study’s outcome measure (the COVID-related non-medical Personal Impact T-Score). Additionally, we used Principal Components Analysis to cluster the fourteen individual impact items, yielding four subscores of non-medical personal impact: Personal Actions, Supply-related, Cancellations and Postponements, and Livelihood. As with the main Impact Scores, these subscores were computed using Streiner et al.’s approach^19^ and their standardized T-score values are used in analysis.

#### Predictor variables

Motivated by the medical ecological framework of this study,^15^ we included a range of variables reflecting sociocultural, environmental, health care, and biological constructs that may be associated with non-medical COVID-19 impacts. This range included demographic variables (race, ethnicity, geographic region of residence, age, gender, sexual orientation, education, religious identification, material assets), psychosocial indices (Perceived Stress Scale, Multidimensional Perceived Social Support Scale, Multidimensional Health Locus of Control Scale), COVID-19-specific variables (COVID-19-related prevention strategies, impact, knowledge, attitudes, institutional effectiveness perception, self-reported COVID infection), social media usage, and health-related variables (difficulty obtaining care, have chronic illness, general health status, work in health care setting).

### Sample Size

The parent study, of which this secondary analysis of non-medical COVID-19-related impact is part, was powered to detect differences in non-medical COVID-19-related impact on lived experience by six world regions. The final sample for the parent study included 10,610 completed surveys from 184 countries across the six global regions. We conducted a post hoc power analysis for this secondary study of non-medical COVID-19 impact; for most variables, the study has >80% power to detect a null difference in proportion of 0.006, assuming a prevalence proportion of 0.01 and α = .05 (JMP Pro 15.0.0 (SAS Institute Inc., Cary, NC)).

### Statistical Analysis

The primary outcome for this study is the non-medical COVID-19 impact dichotomized at the median into two categories: above the median number of items, or at or below the median number of items. We present the previously noted potential predictor variables alongside this outcome variable, showing non-medical COVID-19 impacts with numbers, percentages, and measures of association (chi-square with associated p-value). We use logistic regression to compute odds ratios with 95% confidence intervals for predictor variables in relation to the outcome, to display the magnitude and precision of association.

We aimed to create a parsimonious statistical model to predict the non-medical COVID-19 impact by Deaf and hearing status. We first conducted bivariate analyses to identify associations between variables and Deaf status and non-medical COVID-19 impact, using the Pearson chi-square test as our measure of association. For the overall study, we considered a two-sided asymptotic chi-square p-value < 0.05 to be significant. For our multivariate models, all variables at least marginally associated with Deaf status and non-medical COVID-19 impact were included. We used backward elimination to remove variables from the multivariate model.^20^ We used 0.25 as the cutoff p-value for entry into the model and 0.10 as the threshold for remaining in the model.^21^ We evaluated the model fit using the Pearson chi-square goodness-of-fit test and p-value. An appropriate fit model was defined as one with a p-value greater than 0.05. For bivariate analyses, we used logistic regression-derived Odds Ratios (OR) and their corresponding 95% confidence intervals to denote the magnitude of association. We used adjusted Odds Ratios (aOR) to indicate the same from multivariate analyses. SPSS 29.0.2.0 (IBM Corporation, Armock, NY, USA) was used for analyses.

### Ethical Review

Participants provided informed consent to enroll in this study after reviewing an Information Sheet presented in English, French, Spanish, Italian, Mandarin, Hindi, Arabic, or ASL. Participants were able to skip questions in the survey except age and country of residence. All staff associated with this study completed CITI-Program’s Research, Ethics and Compliance Training. This study was performed in accordance with the ethical standards established by the 1964 Declaration of Helsinki and its later amendments, and adheres to the requirements of the European Union’s General Data Protection Regulation. The University’s IRB determined that this study met federal and University criteria for exemption (Study #00004825).

## Results

Shown in Table 1, the analytic sample comprised 7,998 participants, categorized by hearing status: Deaf (n=304; 3.8%), Hard of Hearing (HoH; n=951; 11.9%), and Hearing (n=6,743; 84.3%). Deaf participants were younger than HoH or hearing counterparts (67.0% aged 18-49 vs. 33.2% HoH, 57.3% hearing; p<0.001) and were more likely to be located in North America (47.7% vs. 27.2% HoH, 19.3% hearing; p<0.001). While 88.0% of Deaf participants had completed at least some college education (vs. 81.7% HoH, 84.9% hearing), postgraduate degree completion was lower in this group. Deaf participants showed higher male representation (51.8%) and distinct language patterns, with 62.2% completing the survey in English only, compared with 71.2% of HoH participants and 57.6% of hearing participants. Approximately 12% of the Deaf participants completed the survey in American Sign Language (ASL) or in the hybrid English version that accompanied the ASL-based sample recruitment. A higher proportion of Deaf participants were included in the first wave of the survey. Across the two administrations of the survey, Deaf participants responded from 54 countries, HoH from 84 countries, and hearing from 184 countries.

**Table 1.** Demographic characteristics by hearing status.

| **Demographic Characteristic** | **Deaf**<br>**(n=304)** | **Hard of**<br>**Hearing**<br>**(n=951)** | **Hearing**<br>**(n=6743)** | **Chi-square**<br>**(p-value)** |
| --- | --- | --- | --- | --- |
| **Age (years)** |  |  |  |  |
| 18-29 | 28.7 (81) | 10.5 (96) | 19.5 (1287) | 416.690 (<0.001) |
| 30-39 | 21.6 (61) | 11.2 (103) | 19.6 (1297) |  |
| 40-49 | 16.7 (47) | 11.6 (106) | 18.2 (1199) |  |
| 50-59 | 13.8 (39) | 18.0 (165) | 19.1 (1262) |  |
| 60-69 | 14.5 (41) | 26.3 (241) | 17.5 (1154) |  |
| 70+ | 4.6 (13) | 22.5 (206) | 6.1 (404) |  |
| Missing (n) | (22) | (40) | (140) |  |
| **Reproductive Age** |  |  |  |  |
| 18-49 | 67.0 (189) | 33.2 (305) | 57.3 (3787) | 206.347 (<0.001) |
| 50+ | 33.0 (93) | 66.8 (618) | 42.7 (2821) |  |
| **World region of residence** |  |  |  |  |
| Africa | 3.9 (12) | 7.8 (74) | 10.2 (686) | 251.676 (p<0.001) |
| Asia | 23.4 (71) | 14.9 (142) | 17.1 (1151) |  |
| Europe | 8.6 (26) | 19.7 (187) | 25.5 (1717) |  |
| Latin America and the Caribbean | 9.9 (30) | 15.7 (149) | 19.4 (1307) |  |
| Northern America | 47.7 (145) | 27.2 (258) | 19.3 (1299) |  |
| Oceania | 6.6 (20) | 14.7 (140) | 8.6 (580) |  |
| Missing (n) | (0) | (1) | (3) |  |
| **Resident of Northern America** |  |  |  |  |
| Yes, Northern America | 47.7 (145) | 27.2 (258) | 19.3 (1299) | 162.386 (<0.001) |
| No, Outside Northern America | 52.3 (159) | 72.8 (692) | 80.7 (5441) |  |
| **Survey Language Completed** |  |  |  |  |
| Arabic |  |  |  | 802.022 (<0.001) |
| American Sign Language (ASL) | 0.0 (0) | 0.8 (8) | 1.8 (123) |  |
| Mandarin (Chinese) | 2.0 (6) | 0.0 (0) | 0.0 (1) |  |
| English | 0.7 (2) | 0.2 (2) | 1.1 (77) |  |
| English/ASL | 62.2 (189) | 71.2 (677) | 57.5 (3881) |  |
| French | 9.9 (30) | 0.7 (7) | 0.1 (4) |  |
| Hindi | 5.6 (17) | 8.7 (82) | 12.6 (847) |  |
| Italian | 1.6 (5) | 0.9 (9) | 1.2 (84) |  |
| Spanish | 8.9 (27) | 4.0 (38) | 6.4 (435) |  |
| Missing (n) | 9.3 (28) | 13.4 (128) | 19.2 (1291) |  |
|  | (0) | (0) | (0) |  |
| **Completed Survey in English** |  |  |  |  |
| Yes, English only |  |  |  | 65.242 (<0.001) |
| No, Other languages (including ASL and English/ASL) | 62.2 (189) | 71.2 (677) | 57.6 (3881) |  |
|  | 37.8 (115) | 28.8 (274) | 42.4 (2862) |  |

**Table 1. Demographic characteristics by hearing status**
| Demographic Characteristic | Deaf<br>(n=304) | Hard of<br>Hearing<br>(n=951) | Hearing<br>(n=6743) | Chi-square<br>(p-value) |
| --- | --- | --- | --- | --- |
| **Survey Wave** |  |  |  |  |
| Early COVID (2020) | 75.0 (228) | 45.6 (434) | 48.6 (3280) | 86.603 (<0.001) |
| Later COVID (2022-23) | 25.0 (76) | 54.4 (517) | 51.4 (3463) |  |
| Missing (n) | (0) | (0) | (0) |  |
| **Gender** |  |  |  |  |
| Male | 51.8 (157) | 50.6 (477) | 45.6 (3061) | 39.503 (<0.001) |
| Female | 44.9 (138) | 48.5 (457) | 53.7 (3602) |  |
| Other gender | 3.3 (10) | 1.0 (9) | 0.7 (45) |  |
| Missing (n) | (1) | (8) | (35) |  |
| **Highest education level completed** |  |  |  |  |
| Did not complete high school | 2.7 (8) | 3.2 (30) | 2.1 (142) | 15.035 <0.058) |
| Completed secondary, high school | 9.2 (27) | 15.1 (140) | 12.9 (856) |  |
| Attended some college | 12.0 (35) | 13.0 (121) | 12.6 (837) |  |
| Graduated college | 42.5 (124) | 39.6 (368) | 43.1 (2853) |  |
| Post-graduate degree | 33.6 (98) | 29.1 (270) | 29.2 (1931) |  |
| Missing (n) | (12) | (22) | (124) |  |
| **Educational level, completed at least some college** |  |  |  |  |
| Yes, completed some college | 88.0 (257) | 81.7 (759) | 84.9 (5621) | 9.141 (0.010) |
| No, completed no college | 12.0 (35) | 18.3 (170) | 15.1 (998) |  |
| **Child care responsibilities** |  |  |  |  |
| Yes | 47.2 (142) | 35.5 (329) | 32.6 (2175) | 13.863 (<0.001) |
| No | 52.8 (159) | 67.7 (634) | 67.4 (4503) |  |
| Missing (n) | (3) | (20) | (116) |  |
| **Elder care responsibilities** |  |  |  |  |
| Yes | 42.5 (128) | 32.3 (302) | 39.3 (2603) | 13.125 (0.001) |
| No | 57.5 (173) | 64.7 (602) | 60.7 (4024) |  |
| Missing (n) | (3) | (15) | (65) |  |
| **Own Home** |  |  |  |  |
| Yes | 70.0 (205) | 74.0 (687) | 68.0 (4488) | 13.819 (<001) |
| No/Don't know | 30.0 (88) | 26.0 (242) | 32.0 (2116) |  |
| Missing (n) | (11) | (22) | (139) |  |
| **Own Car** |  |  |  |  |
| Yes | 68.2 (202) | 75.5 (707) | 66.9 (4454) | 28.195 (<001) |
| No/Don't know | 31.8 (94) | 24.9 (229) | 33.1 (2205) |  |
| Missing (n) | (8) | (15) | (84) |  |

Table 2 presents psychosocial characteristics by hearing status. Deaf participants showed stronger “external” health locus of control dimensions: 61.6% scored above the median on the “Chance” subscale (vs. 39.0% HoH, 35.9% hearing participants; p<0.001) and 47.0% relied on “Powerful Others” that control their health (vs. 39.2% HoH, 35.6% hearing; p<0.001). Perceived social support was significantly lower among Deaf participants, with 38.4% in the lowest quartile compared to 28.6% of HoH and 24.1% of hearing participants (p<0.001). Overall, 12.8% of Deaf participants reported the highest level of social support, versus 25.9% of hearing participants. COVID-19-related stigma was significantly higher among Deaf participants: 35.5% believed that individuals infected with COVID-19 would lose community respect or status (vs. 21.1% HoH, 16.8% hearing; p<0.001), and 41.9% of Deaf participants reported that people gossip or think negatively about those suspected or have coronavirus (vs. 34.3% HoH, 33.2% hearing; p<0.001), and 20.5% reported experiencing coronavirus-related harassment or bullying (vs. 19.4% HoH, 15.2% hearing; p<0.001).

**Table 2.** Psychosocial variables by hearing status.

| Psychosocial variable | Deaf<br>(n=304) | Hard of<br>Hearing<br>(n=951) | Hearing<br>(n=6743) | Chi-square<br>(p-value) |
| --- | --- | --- | --- | --- |
| **Multidimensional Health Locus of Control – Internal “Internality” subscale** |  |  |  |  |
| Stronger (above median) | 30.1 (89) | 26.6 (247) | 30.7 (2029) | 6.640 (0.036) |
| Weaker (at or below median) | 69.9 (207) | 73.4 (682) | 69.3 (4573) |  |
| Missing (n) | (8) | (22) | (141) |  |
| **Multidimensional Health Locus of Control – Powerful Others subscale** |  |  |  |  |
| Stronger (above median) | 47.0 (139) | 39.2 (362) | 35.6 (2353) | 19.078 (<0.001) |
| Weaker (at or below median) | 53.0 (157) | 60.8 (561) | 64.4 (4251) |  |
| Missing (n) | (8) | (28) | (139) |  |
| **Multidimensional Health Locus of Control – Chance (externality) subscale** |  |  |  |  |
| Stronger (above median) | 61.6 (183) | 39.0 (357) | 35.9 (2365) | 81.852 (<0.001) |
| Weaker (at or below median) | 38.4 (114) | 61.0 (559) | 64.1 (4225) |  |
| Missing (n) | (7) | (35) | (153) |  |
| **Perceived Social Support Scale - Total** |  |  |  |  |
| Lowest social support (1<sup>st</sup> quartile) | 38.4 (114) | 28.6 (247) | 24.1 (1534) | 61.008 (<0.001) |
| Low-Moderate social support (2<sup>nd</sup> quartile) | 29.4 (85) | 27.9 (241) | 26.0 (1655) |  |
| Moderate-High social support (3<sup>rd</sup> quartile) | 18.3 (53) | 19.4 (168) | 23.9 (1520) |  |
| Highest social support (4<sup>th</sup> quartile) | 12.8 (37) | 24.1 (208) | 25.9 (1647) |  |
| Missing (n) | (15) | (87) | (387) |  |
| **Do people talk badly or gossip about other people who are living with, have had, or are thought to have coronavirus?** |  |  |  |  |
| Definitely/Probably Yes | 41.9 (126) | 34.3 (324) | 33.2 (2222) | 9.989 (0.007) |
| Definitely/Probably No, or Don't Know | 58.1 (175) | 65.7 (620) | 66.8 (4478) |  |
| Missing (n) | (3) | (7) | (43) |  |
| **Do people who have had coronavirus infection (or COVID-19) lose respect or status in the community?** |  |  |  |  |
| Definitely/Probably Yes | 35.5 (108) | 21.1 (200) | 16.8 (1130) | 75.681 (<0.001) |
| Definitely/Probably No, or Don't Know | 64.5 (196) | 78.9 (748) | 83.2 (5590) |  |
| Missing (n) | (0) | (3) | (23) |  |
| **Have you or a family member been harassed, bullied, or hurt because of coronavirus?** |  |  |  |  |
| Yes | 20.5 (62) | 19.4 (183) | 15.2 (1018) | 15.883 (<0.001) |
| No/DK | 79.5 (261) | 80.6 (182) | 84.8 (5692) |  |
| Missing | (1) | (6) | (33) |  |

Most non-medical impacts of the pandemic were experienced significantly more frequently among Deaf participants versus their HoH and hearing counterparts across employment, resource access, and healthcare domains (Table 3). Deaf participants reported higher rates of stockpiling food and household supplies (70.4% vs 59.2% HoH, 60.2% hearing, p<0.001) and obtaining extra prescription medication refills (49.7% vs 30.8% HoH, 28.0% hearing, p<0.001), alongside greater inability to obtain groceries (28.6% vs. 22.3% HoH, 20.6% hearing; p=0.002), cleaning supplies (37.3% vs. 26.6% HoH, 25.1% hearing; p<0.001), and prescription medications (17.2% vs. 13.1% HoH,10.3% hearing; p<0.001). Employment-related impacts were significantly more common among Deaf participants: 28.9% quit jobs due to the pandemic, compared with 13.2% of HoH participants and 9.6% of hearing participants (p<0.001). Healthcare access was significantly disrupted, with 39.9% of Deaf participants postponing or canceling medical procedures or surgery, compared with 31.7% of HoH and 24.6% of hearing participants (p<0.001). In total, 55.9% of Deaf participants scored above the median on the composite non-medical impact index (vs 41.2% HoH, 39.2% hearing; p<0.001), with 41.8% of Deaf participants scoring in the highest quartile (vs. 24.8% HoH, 23.9% hearing; p<0.001). No significant difference was noted in non-medical impact between HoH v. hearing (p=0.220; data not shown).

**Table 3.** Non-medical COVID impact by hearing status.

| **Health Outcomes** | **Deaf**<br>**(n=304)** | **Hard of**<br>**Hearing**<br>**(n=951)** | **Hearing**<br>**(n=6743)** | **Chi-square**<br>**(p-value)** |
| --- | --- | --- | --- | --- |
| **Decided not to travel or changed travel plans because of coronavirus outbreak** |  |  |  |  |
| Yes | 81.5 (247) | 70.8 (667) | 73.3 (4909) | 13.453 (<0.001) |
| No/Don't know | 18.5 (56) | 29.2 (275) | 26.7 (1788) |  |
| Missing (n) | (1) | (9) | (46) |  |
| **Stocked up on items such as food and household supplies because of coronavirus outbreak** |  |  |  |  |
| Yes | 70.4 (212) | 59.2 (557) | 60.2 (4018) | 13.350 (<001) |
| No/Don't know | 29.6 (89) | 40.8 (384) | 39.8 (2657) |  |
| Missing (n) | (3) | (10) | (68) |  |
| **Postponed or canceled health care visits because of coronavirus outbreak** |  |  |  |  |
| Yes | 54.1 (164) | 49.3 (463) | 48.7 (3261) | 3.438 (0.79) |
| No/Don't know | 45.9 (139) | 50.7 (476) | 51.3 (3433) |  |
| Missing (n) | (1) | (12) | (49) |  |
| **Got extra refills on prescription medication because of coronavirus outbreak** |  |  |  |  |
| Yes | 49.7 (149) | 30.8 (287) | 28.0 (1869) | 66.306 (<0.001) |
| No/Don't know | 50.3 (151) | 69.2 (645) | 72.0 (4797) |  |
| Missing (n) | (4) | (19) | (77) |  |
| **Stayed home instead of going to work school, or other regular activities because of coronavirus outbreak** |  |  |  |  |
| Yes | 74.2 (224) | 68.7 (644) | 71.2 (4755) | 4.045 (0.132) |
| No/Don't know | 25.8 (78) | 31.3 (294) | 28.8 (1928) |  |
| Missing (n) | (2) | (13) | (60) |  |
| **Postponed or canceled a medical procedure or surgery because of coronavirus outbreak** |  |  |  |  |
| Yes | 39.9 (121) | 31.7 (296) | 24.6 (1638) | 53.192 (<0.001) |
| No/Don't know | 60.1 (182) | 68.3 (638) | 75.4 (5024) |  |
| Missing (n) | (1) | (17) | (81) |  |

**Table 3. Non-medical COVID impact by hearing status**
| **Health Outcomes** | **Deaf**<br>**(n=304)** | **Hard of**<br>**Hearing**<br>**(n=951)** | **Hearing**<br>**(n=6743)** | **Chi-square**<br>**(p-value)** |
| --- | --- | --- | --- | --- |
| **Canceled plans to attend large gatherings such as concerts or sporting events because of coronavirus outbreak** |  |  |  |  |
| Yes | 71.7 (218) | 67.5 (629) | 71.0 (4739) | 5.141 (0.076) |
| No/Don't know | 28.3 (86) | 32.5 (303) | 29.0 (1932) |  |
| Missing (n) | (0) | (19) | (69) |  |
| **Quit my job because of coronavirus outbreak** |  |  |  |  |
| Yes | 28.9 (86) | 13.2 (121) | 9.6 (632) | 117.209 (<0.001) |
| No/Don't know | 71.1 (212) | 86.8 (798) | 90.4 (5971) |  |
| Missing (n) | (6) | (32) | (140) |  |
| **Lost income from a job or business because of coronavirus outbreak** |  |  |  |  |
| Yes | 40.7 (123) | 35.6 (337) | 38.0 (2551) | 3.213 (0.201) |
| No/Don't know | 59.3 (179) | 64.4 (610) | 62.0 (4158) |  |
| Missing (n) | (2) | (4) | (34) |  |
| **Been unable to get groceries because of coronavirus outbreak** |  |  |  |  |
| Yes | 28.6 (87) | 22.3 (210) | 20.6 (1379) | 12.087 (0.002) |
| No/Don't know | 71.4 (217) | 77.7 (731) | 79.4 (5310) |  |
| Missing (n) | (0) | (10) | (54) |  |
| **Been unable to get cleaning supplies or hand sanitizer because of coronavirus outbreak** |  |  |  |  |
| Yes | 37.3 (112) | 26.6 (249) | 25.1 (1680) | 22.927 (<0.001) |
| No/Don't know | 62.7 (188) | 73.4 (687) | 74.9 (5015) |  |
| Missing (n) | (4) | (15) | (48) |  |
| **Been unable to get prescription medication because of coronavirus outbreak** |  |  |  |  |
| Yes | 17.2 (52) | 13.1 (123) | 10.3 (690) | 19.391 (<0.001) |
| No/Don't know | 82.8 (250) | 86.9 (813) | 89.7 (5986) |  |
| Missing (n) | (2) | (15) | (67) |  |
| **COVID non-medical impact index, by quartile** |  |  |  |  |
| Low number of items (Q1) | 10.2 (31) | 18.6 (176) | 17.6 (1184) | 54.624 (<0.001) |
| Low-moderate number of items (Q2) | 21.4 (65) | 26.5 (251) | 28.2 (1894) |  |
| Moderate-high number of items (Q3) | 26.6 (81) | 30.2 (286) | 30.4 (2043) |  |
| High number of items (Q4) | 41.8 (127) | 24.8 (235) | 23.9 (1604) |  |
| Missing (n) | (0) | (3) | (18) |  |

**Table 3. Non-medical COVID impact by hearing status**
| **Health Outcomes** | **Deaf**<br>**(n=304)** | **Hard of**<br>**Hearing**<br>**(n=951)** | **Hearing**<br>**(n=6743)** | **Chi-square**<br>**(p-value)** |
| --- | --- | --- | --- | --- |
| **COVID non-medical impact index, by**<br>**median** |  |  |  |  |
| Higher impact (above median) | 55.9 (170) | 41.2 (391) | 39.2 (2634) | 34.638 (<0.001) |
| Lower impact (at or below median) | 44.1 (134) | 58.8 (557) | 60.8 (4091) |  |
| Missing (n) | (0) | (3) | (18) |  |

In the past 30 days, shown in Table 4, 79.4% of Deaf participants experienced poor mental health (vs. 65.8% HoH, 64.0% hearing; p<0.001), 73.1% reported poor physical health (vs. 63.6% HoH, 51.5% hearing; p<0.001), and 70.8% had limitations on their usual activities due to poor physical and mental health (vs. 52.5% HoH, 45.7% hearing; p<0.001). Cost-related healthcare access barriers were nearly twice as prevalent among Deaf participants (38.5% vs. 28% HoH, 21.6% hearing; p<0.001). HoH participants showed poorer general health ratings, with 20.4% reporting fair or poor health compared to 13.5% of Deaf participants and 13.3% of hearing participants (p<0.001). Self-reported coronavirus infection rates were elevated among both Deaf (17.9%) and HoH (17.5%) participants compared to hearing participants (12.0%; p<0.001).

**Table 4.** Health-related COVID variables by hearing status.

| **Health-related variable** | **Deaf**<br>**(n=304)** | **Hard of**<br>**Hearing**<br>**(n=951)** | **Hearing**<br>**(n=6743)** | **Chi-square**<br>**(p-value)** |
| --- | --- | --- | --- | --- |
| **Do you feel that worry or stress related to coronavirus has had a negative impact on your mental health, or not?** |  |  |  |  |
| Yes, major or minor impact | 67.0 (203) | 66.0 (626) | 66.5 (4471) | 0.139 (0.933) |
| No/Don't know | 33.0 (100) | 34.0 (323) | 33.5 (2256) |  |
| Missing (n) | (1) | (2) | (6) |  |
| **Do you feel you now have - or have you recently had - coronavirus infection?** |  |  |  |  |
| Yes have/had coronavirus | 17.9 (54) | 17.5 (165) | 12.0 (799) | 30.130 (p<0.001) |
| No/Don't know | 82.1 (247) | 82.5 (777) | 88.0 (5887) |  |
| Missing (n) | (3) | (9) | (57) |  |
| **Was there a time in the past 12 months when you needed to get health care (for example, see a doctor), but could not because of cost?** |  |  |  |  |
| Yes | 38.5 (115) | 28.0 (264) | 21.6 (1448) | 60.567 (p<0.001) |
| No/Don't know | 61.5 (184) | 72.0 (679) | 78.4 (5243) |  |
| Missing (n) | (5) | (8) | (52) |  |
| **Would you say in general your health is:** |  |  |  |  |
| Fair/poor | 13.5 (41) | 20.4 (193) | 13.3 (895) | 34.092 (<0.001) |
| Excellent/Very good/Good | 86.5 (262) | 79.6 (753) | 86.7 (5815) |  |
| Missing (n) | (1) | (5) | (33) |  |
| **Thinking about your physical health, which includes physical illness and injury, for how many days during the past 30 days was your physical health not good?** |  |  |  |  |
| One or more days | 73.1 (193) | 63.6 (564) | 51.5 (3275) | 87.747 (<0.001) |
| No days | 26.9 (71) | 36.4 (323) | 48.5 (3090) |  |
| Missing (n) | (40) | (64) | (378) |  |
| **Thinking about your mental health, which includes stress, depression, and problems with emotions, for how many days during the past 30 days was your mental health not good?** |  |  |  |  |
| One or more days | 79.4 (212) | 65.8 (577) | 64.0 (4082) | 27.096 (<0.001) |
| No days | 20.6 (55) | 34.2 (300) | 36.0 (2296) |  |
|  | (37) | (74) | (365) |  |

**Table 4. Health-related COVID variables by hearing status**
| **Health-related variable** | **Deaf**<br>**(n=304)** | **Hard of**<br>**Hearing**<br>**(n=951)** | **Hearing**<br>**(n=6743)** | **Chi-square**<br>**(p-value)** |
| --- | --- | --- | --- | --- |
| Missing (n) |  |  |  |  |
| **During the past 30 days, for about**<br>**how many days did poor physical or**<br>**mental health keep you from doing**<br>**your usual activities, such as self-**<br>**care, work, or recreation?** |  |  |  |  |
| One or more days | 70.8 (189) | 52.5 (461) | 45.7 (2901) | 75.050 (<0.001) |
| No days | 29.2 (78) | 47.5 (417) | 54.3 (3446) |  |
| Missing (n) | (37) | (73) | (396) |  |

Multinomial logistic regression examined associations between hearing status and high non-medical COVID-19 impact across three progressively adjusted models (Table 5). In Model 1 (unadjusted), Deaf participants had twice the odds of high non-medical COVID-19 impact compared with hearing participants (OR=2.0, 95% CI: 1.6-2.5, p<0.001), which demonstrates the baseline disparity magnitude. Model 2 adjusted for demographics and structural determinants (survey wave, education status, child/elder caregiving responsibilities, car ownership, and health locus of control), reducing the odds ratio to 1.7 (95% CI: 1.2-2.5, p<0.001). Model 3 incorporated potential mediators (inability to obtain healthcare, bullying experiences, COVID-related stigma, HRQOL days lost, general self-reported health status, and COVID-related worry), progressively reducing the odds ratio to 1.6 (95%:1.1-2.4, p<0.001). Deaf status remains a statistically significant independent predictor of higher non-medical COVID-19 impact than hearing participants. Furthermore, HoH status showed no statistically significant differences compared with their hearing counterparts in unadjusted or adjusted models.

**Table 5.** Hearing Status and Non-Medical COVID-related Impact, Multivariate multinomial logistic regression results.

| **Model** | **Adjustment**<br>(variables remaining in model) | **aOR (95% CI)**<br>**Deaf vs**<br>**Hearing** | **aOR (95% CI)**<br>**HoH vs**<br>**Hearing** | **Chi-square**<br>**(p-value)** |
| --- | --- | --- | --- | --- |
| **Model 1** | Hearing status only | 2.0 (1.6, 2.5) | 1.1 (0.9, 1.3) | 34.638<br>(<0.001) |
| **Model 2** | + Demographics (variables remaining significant in the model: survey wave, education, child/elder care, car ownership, MHLC) * | 1.7 (1.2, 2.5) | 1.1 (0.9, 1.3) | 271.141<br>(<0.001) |
| **Model 3** | + Mediators (variables remaining significant in the model: Could not obtain health care, bullied, gossip/lose respect for people with COVID, HRQOL days lost, general self-report health status, COVID worry index) * | 1.6 (1.1, 2.4) | 1.0 (0.8, 1.3) | 685.976<br>(<0.001) |

## Discussion

We demonstrated that Deaf people around the world disproportionately experienced greater non-medical COVID-19 impact compared with people who are Hard of Hearing or with people who are hearing, independent of demographic characteristics, psychosocial pathways, and health status. Importantly, the Deaf community’s risks were distinct and often significantly different from those who are HoH on most variables, underscoring the importance of analyzing Deaf and HoH statuses separately. In addition, this suggests that the elevated risk of non-medical COVID-19 impact is largely due to Deaf status itself associated with sign language access and Deaf cultural identity, not due to hearing loss per se. This aligns with the increasingly growing understanding that Deaf people and communities are also a sociolinguistic minority with socially constructed identities, not just a group identified solely by disability.^22,23^

This persistently significant predictor of Deaf status after adjustment points to structural inequalities, mainly audism and system-wide barriers that drive these disparities. Audism, discrimination and prejudice against deaf people rooted in the belief that one is superior because of one’s ability to hear and speak, manifests at both the individual and institutional levels of oppression.^24,25^ Deaf people experience marginalization of their identities and languages when navigating hearing environments that are not designed in their favor. Healthcare systems are influenced by audist assumptions which lead to inadequate, unequal access to accommodations and inclusion for Deaf people.^26,27^ Several system-wide barriers include inadequate access to communication, accommodation failures, an external locus of control as a structural adaptation, increased stigma and social isolation, and disruptions in access to resources, employment, and healthcare.

Because the Deaf community relies on visual language that their families are typically not fluent in, inadequate language access during childhood is associated with persistent adverse outcomes into adulthood.^28^ During the COVID-19 pandemic, Deaf individuals experienced disproportionate barriers to timely access to critical information and resources, exposing longstanding inequities in communication access. Language deprivation, the denial of full access to a spoken language and/or natural sign language during critical development periods, exacerbated health disparities through reduced health literacy and phenomena such as “Dinner Table Syndrome,” which reflects chronic exclusion from incidental social learning.^29,30^ Pandemic-specific communication practices, including face mask mandates that obscured facial expressions essential for visual communication, social distancing that prevented proximity-dependent communication techniques, widespread shift to audio-only virtual formats (without captioning or sign language interpretation), and inaccessible public health messaging, further excluded Deaf individuals from essential health information. The rapid shift to remote services during COVID further widened accessibility gaps, limiting Deaf people’s ability to access necessary supports.

Deaf participants exhibited significantly higher external locus of control over health outcomes, attributing them to “Chance” and “Powerful Others.” This heightened perception of an external locus of control reflects institutional obstacles to accessing healthcare, obtaining timely health information, communicating, and receiving appropriate accommodations. When Deaf individuals attempt to navigate systems that fail to meet their needs, they may feel they have no choice but to adapt to systemic limitations. Perceived social support was diminished among Deaf participants, reflecting social isolation that may have been exacerbated by social distancing requirements and mask mandates that obscured facial expressions critical to visual communication. Also, disruptions to Deaf community gatherings and signing spaces may have contributed to greater isolation and less health information exchange. The higher level of COVID-19-related stigma reflects vulnerabilities that Deaf people face in hearing majority environments that include intersectional identities. External locus of control, diminished social support, and heightened stigma exposure represent psychosocial factors influencing health disparities.

Pronounced disruptions in obtaining essential resources, such as higher rates of stockpiling behavior and obtaining extra prescription medication refills, show anticipatory responses to anticipated access barriers. Deaf individuals reported greater difficulty obtaining groceries, cleaning supplies, and prescription medications, underscoring structural barriers in basic survival and chronic disease management during public health crises. Employment disruptions were affected by systemic workplace barriers since a higher percentage of Deaf people reported quitting their jobs compared to hearing peers, and yet, overall income loss did not differ across all groups. This suggests that job termination, rather than reduced work hours, drove employment-related impacts. These patterns also could reflect communication barriers in the workplace, accommodation failures, and/or hostile work environments in which Deaf individuals were unable to function in pandemic conditions. The rapid move to remote work without adequate accessibility accommodations may have further intensified these barriers.

Healthcare-related access was significantly higher for Deaf participants, suggesting that employment disruption and related challenges may have contributed to poorer health outcomes. Deaf individuals reported worse mental and physical health and greater limitations in daily activities due to poor health, compared to hearing participants. Increased coronavirus infection rates among Deaf individuals may be attributable to delayed or inaccessible public health information and persistent communication barriers. Although COVID-19-related worry was similar across all groups, disparities in mental health burden, physical health limitations, and healthcare costs were more pronounced among Deaf participants, indicating that system-wide barriers, rather than individual risk perception, drove health inequities. HoH individuals also showed poorer health outcomes, a unique finding in this population, underscoring the need for greater attention to health navigation difficulties across language modalities and degrees of hearing loss. Altogether, the data point to a cumulative burden determined by language modality and access, with Deaf sign language users facing distinct and compounding structural disadvantages.

Health equity efforts designed to achieve true cultural competency include moving beyond individualized approaches and tackling structural barriers faced by Deaf communities. Primary recommendations include adopting universal design principles throughout healthcare and public services, reducing the “Deaf Tax” associated with repeatedly requesting accommodations,^31^ formally recognizing sign languages as fully developed languages that require protection, and centering Deaf knowledge, leadership, and lived experience in research, policy development, and emergency preparedness.

Most health equity frameworks focus on individual behaviors. Deafness is a structural determinant of health, comparable to race, socioeconomic status, and gender, as a fundamental cause of health inequities.^32^ Efficient interventions must focus on system-wide barriers rather than individual deficits. These include enforcing accessibility policies, adopting inclusive technologies, providing Deaf cultural competency training for healthcare providers, dismantling audist institutional practices, and integrating accessible communication (signed, captioned, and interpreted modalities) as a core foundation of healthcare rather than optional accommodations.

Our study has several limitations. The focus on pandemic-related, non-medical COVID-19 impacts limits causal inference and fails to capture longer-term changes in mental and physical health or access barriers beyond the acute pandemic period. Language access in research for Deaf people is challenging and although we offered a fully ASL-deployed version during the second wave of our study, it was not used frequently to complete the survey. We did not offer the survey in other signed languages during either wave. The strengths of our study included the broad geographic region and the written languages we included, which together created a large global sample. Additionally, we distinctly separated people who self-identify as “Deaf” from those who self-identify as “Hard of Hearing,” rather than aggregating the two populations. This disaggregation is rare in research on Deaf communities, though numerous studies have shown that these populations are distinct and face unique barriers.^33–36^ One qualitative study based in the U.S. found that Deaf patients relied heavily on ASL while HoH patients relied on lipreading and written notes, demonstrating distinct cultural identities and communication needs that are not always recognized by healthcare providers.^33^ Analyzing these populations separately allowed us to better understand variations in experience and impact.

Longitudinal research is needed to examine how access to communication and institutional responses shape health trajectories throughout time. While multiple demographic and socioeconomic factors were accounted for in this study, the full effects of audism, accommodation failures, and discrimination across healthcare, employment, and social systems were not fully captured. Qualitative studies that examine the lived experiences of Deaf individuals’ communication access and exclusion from information networks are needed to explain the persistent risk of Deaf status. Deaf and HoH populations differ in language modalities, hearing loss, and cultural affiliations. The absence of high risk among HoH participants shows that cultural and linguistic aspects, rather than hearing loss alone, drive the observed disparities. This warrants further investigation throughout healthcare systems and geographic contexts.

## Conclusion

This study shows that Deafness functions as a structural determinant of health inequity, with disproportionate non-medical impacts during the COVID-19 pandemic driven by language access barriers and institutional exclusion rather than hearing loss itself. Responding to these inequities requires systemic public health interventions that include accessible communication, the dismantling of audist policies, and the centering of Deaf communities in public health preparedness and crisis response.

## Data Availability

All data produced in the present study are available upon reasonable request to the authors

## References

1. Gutierrez-Sigut E, Lamarche VM, Rowley K, et al. How do face masks impact communication amongst deaf/HoH people? Cogn Res Princ Implic. Sep 5 2022;7(1):81. doi:10.1186/s41235-022-00431-4

2. Giovanelli E, Gianfreda G, Gessa E, et al. The effect of face masks on sign language comprehension: performance and metacognitive dimensions. Conscious Cogn. Mar 2023;109:103490. doi:10.1016/j.concog.2023.103490

3. Panko TL, Contreras J, Postl D, et al. The Deaf Community’s Experiences Navigating COVID-19 Pandemic Information. Health Lit Res Pract. Apr 2021;5(2):e162–e170. doi:10.3928/24748307-20210503-01

4. James TG, Helm KVT, Ratakonda S, Smith LD, Mitra M, McKee MM. Health Literacy and Difficulty Accessing Information About the COVID-19 Pandemic Among Parents Who Are Deaf and Hard-of-Hearing. Health Lit Res Pract. Oct 2022;6(4):e310–e315. doi:10.3928/24748307-20221116-01

5. Almusawi H, Alasim K, BinAli S, Alherz M. Disparities in health literacy during the COVID-19 pandemic between the hearing and deaf communities. Res Dev Disabil. Dec 2021;119:104089. doi:10.1016/j.ridd.2021.104089

6. Xu D, Yan C, Zhao Z, Weng J, Ma S. External Communication Barriers among Elderly Deaf and Hard of Hearing People in China during the COVID-19 Pandemic Emergency Isolation: A Qualitative Study. Int J Environ Res Public Health. Nov 2 2021;18(21)doi:10.3390/ijerph182111519

7. Tomasuolo E, Gulli T, Volterra V, Fontana S. The Italian Deaf Community at the Time of Coronavirus. Front Sociol. 2020;5:612559. doi:10.3389/fsoc.2020.612559

8. Engelman A, Paludneviciene R, Wagner K, Jacobs K, Kushalnagar P. Food Worry in the Deaf and Hard-of-Hearing Population During the COVID-19 Pandemic. Public Health Rep. Mar–Apr 2021;136(2):239–244. doi:10.1177/0033354920974666

9. Gillespie AN, Smith L, Shepherd DA, Xu J, Khanal R, Sung V. Socio-Emotional Experiences and Wellbeing of Deaf and Hard of Hearing Children and Their Parents before and during the COVID-19 Pandemic. Children (Basel). Jun 30 2023;10(7)doi:10.3390/children10071147

10. Moreland CJ, Rao SR, Jacobs K, Kushalnagar P. Equitable Access to Telehealth and Other Services for Deaf People During the COVID-19 Pandemic. Health Equity. 2023;7(1):126–136. doi:10.1089/heq.2022.0115

11. Roguski M, Officer TN, Nazari Orakani S, Good G, Handler-Schuster D, McBride-Henry K. Ableism, Human Rights, and the COVID-19 Pandemic: Healthcare-Related Barriers Experienced by Deaf People in Aotearoa New Zealand. Int J Environ Res Public Health. Dec 18 2022;19(24)doi:10.3390/ijerph192417007

12. Turner K, Nguyen OT, Alishahi Tabriz A, Islam JY, Hong YR. COVID-19 Vaccination Rates Among US Adults With Vision or Hearing Disabilities. JAMA Ophthalmol. Sep 1 2022;140(9):894–899. doi:10.1001/jamaophthalmol.2022.3041

13. Nshimirimana DA, Kokonya D, Gitaka J, Wesonga B, Mativo JN, Rukanikigitero JMV. Impact of COVID-19 on health-related quality of life in the general population: A systematic review and meta-analysis. PLOS Glob Public Health. 2023;3(10):e0002137. doi:10.1371/journal.pgph.0002137

14. Clemente-Suárez VJ, Navarro-Jiménez E, Moreno-Luna L, et al. The Impact of the COVID-19 Pandemic on Social, Health, and Economy. Sustainability. 2021;13(11):6314.

15. De Ver Dye T, Muir E, Farovitch L, Siddiqi S, Sharma S. Critical medical ecology and SARS-COV-2 in the urban environment: a pragmatic, dynamic approach to explaining and planning for research and practice. Infectious Diseases of Poverty. 2020;9(1):1–7.

16. von Elm E, Altman DG, Egger M, Pocock SJ, Gøtzsche PC, Vandenbroucke JP. The Strengthening the Reporting of Observational Studies in Epidemiology (STROBE) statement: guidelines for reporting observational studies. The Lancet. 2007;370(9596):1453–1457. doi:10.1016/S0140-6736(07)61602-X

17. Dye T, Levandowski B, Siddiqi S, et al. Non-medical COVID-19-related personal impact in medical ecological perspective: A global multileveled, mixed method study. medRxiv. 2021:2020.12. 26.20248865.

18. Padden CA, Humphries T. Deaf in America. Harvard University Press; 1988.

19. Streiner DL, Norman GR, Cairney J. Health measurement scales: a practical guide to their development and use. Oxford University Press, USA; 2015.

20. Wang X, Cheng Z. Cross-sectional studies: strengths, weaknesses, and recommendations. Chest. 2020;158(1):S65–S71.

21. Bursac Z, Gauss CH, Williams DK, Hosmer DW. Purposeful selection of variables in logistic regression. Source code for biology and medicine. 2008;3(1):1–8.

22. Kelly RR, Hauser PC, Berent GP, Rizzo S, Contreras J, Jamieson JP. Stereotype threat effects on deaf and hard-of-hearing college students’ mathematics performance. J Deaf Stud Deaf Educ. Jan 28 2026;doi:10.1093/jdsade/enaf088

23. Ladd P, Lane, H. Deaf Ethnicity, Deafhood, and Their Relationship. Sign Language Studies. Summer 2013 2013;13(4):565–579. doi:10.1353/sls.2013.0012

24. Humphries T. Communicating Across Cultures (Deaf-Hearing) and Language Learning. Ph.D. Dissertation. Union Graduate School; 1977.

25. Bauman HD. Audism: exploring the metaphysics of oppression. J Deaf Stud Deaf Educ. 2004 Spring 2004;9(2):239–46. doi:10.1093/deafed/enh025 10.1093/deafed/enh025.

26. Eckert RC, Rowley AJ. Audism:A Theory and Practice of Audiocentric Privilege. Humanity & Society. 2013;37(2):101–130. doi:10.1177/0160597613481731

27. Alhasawi Y. Audism: A review. Gallaudet Chronicles of Psychology. 2016;4(1):26–30.

28. Hall WC. What you don’t know can hurt you: The risk of language deprivation by impairing sign language development in deaf children. Maternal and child health journal. 2017;21(5):961–965.

29. Hall WC, Smith SR, Sutter EJ, DeWindt LA, Dye TD. Considering parental hearing status as a social determinant of deaf population health: Insights from experiences of the “dinner table syndrome”. PloS one. 2018;13(9):e0202169.

30. Meek DR. Dinner table syndrome: A phenomonological study of deaf individuals’ experiences with inaccessible communication. The Qualitative Report. 2020;25(6):1676–1694.

31. Aldalur A, Hall WC, DeAndrea-Lazarus IA. No Taxation Without Representation: Addressing the “Deaf Tax” in Academic Medicine. Acad Med. Aug 1 2022;97(8):1123–1127. doi:10.1097/acm.0000000000004685

32. Link BG, Phelan J. Social conditions as fundamental causes of disease. J Health Soc Behav. 1995;Spec No:80–94.

33. Iezzoni LI, O’Day BL, Killeen M, Harker H. Communicating about health care: observations from persons who are deaf or hard of hearing. Ann Intern Med. Mar 2 2004;140(5):356–62. doi:10.7326/0003-4819-140-5-200403020-00011

34. McKee MM, Barnett SL, Block RC, Pearson TA. Impact of communication on preventive services among deaf American Sign Language users. Am J Prev Med. Jul 2011;41(1):75–9. doi:10.1016/j.amepre.2011.03.004

35. Jacob SA, Palanisamy UD, Napier J, Verstegen D, Dhanoa A, Chong EY. Health Care Needs of Deaf Signers: The Case for Culturally Competent Health Care Providers. Acad Med. Mar 1 2022;97(3):335–340. doi:10.1097/ACM.0000000000004181

36. Barnett S, McKee M, Smith SR, Pearson TA. Deaf sign language users, health inequities, and public health: opportunity for social justice. Prev Chronic Dis. Mar 2011;8(2):A45.

